# Results dissemination from completed clinical trials conducted at German university medical centers remains delayed and incomplete. The 2014-2017 cohort

**DOI:** 10.1101/2021.08.05.21261624

**Authors:** Nico Riedel, Susanne Wieschowski, Till Bruckner, Martin R. Holst, Hannes Kahrass, Edris Nury, Joerg J. Meerpohl, Maia Salholz-Hillel, Daniel Strech

**Affiliations:** QUEST Center for Transforming Biomedical Research, Berlin Institute of Health (BIH) at Charité – Universitätsmedizin Berlin, Berlin, Germany; Institute for Ethics, History, and Philosophy of Medicine, Hannover Medical School, Hannover, Germany; Institute for Evidence in Medicine, Medical Center – University of Freiburg, Faculty of Medicine, University of Freiburg, Freiburg, Germany; Cochrane Germany, Cochrane Germany Foundation, Freiburg, Germany; Charité Universitätsmedizin Berlin, Berlin, Germany

## Abstract

**Objective:** Timely publication of clinical trial results is central for evidence-based medicine. The performance of university medical centers (UMCs) regarding timely results publication informs the local and national decision makers, patients, and the public about the need to improve trial results dissemination.

**Study Design and Setting:** Following the same search and tracking methods used in our previous study for the years 2009 - 2013, we identified trials led by German UMCs completed between 2014 and 2017 and tracked results dissemination for the identified trials.

**Results:** We identified 1,658 trials in the 2014-2017 cohort. Of these trials, 43% published results as either journal publication or summary results within 24 months after completion date, which is an improvement of 3.8% percentage points compared to the previous study. At the UMC level, the 24-month publication rates varied from 14% to 71%. Five years after completion, still 30% of the trials remained unpublished.

**Conclusion:** Despite minor improvements compared to the previously investigated cohort, the rate of timely reporting of trials led by German UMCs remains low. German UMCs should take further steps to improve timely reporting rates.

## Background

Timely and comprehensive reporting of clinical trial results is a crucial aspect of evidence-based health care and responsible research. In a previous study, we tracked results reporting in journals and registries for all registered trials from German University Medical Centers (UMCs) that were completed between 2009 and 2013 [1]. Its results were also presented via an interactive website benchmarking German UMCs on their performance in results dissemination [2].

In that former study, we identified 2,132 clinical trials via the registries ClinicalTrials.gov (n=1,905) and the German Clinical Trials Register (DRKS) (n=227) that i) recruited trial participants at one or more German UMCs and ii) had their primary completion date (last visit of last patient for a primary outcome measure) between 2009 and 2013. These trials included 506,876 anticipated participants. The study found that only 39% of all registered clinical trials conducted at one of the 36 German UMCs published their findings as either registry summary results or a journal publication in a timely manner within 24 months after the trial’s completion date (last visit of last patient for primary and secondary outcome measures and adverse events). Five years after the completion date, 67% of all trials had published their results. In other words: applying extensive search strategies to identify trial publications, it appears that five years after study completion 33% of all trials remained unpublished.

The study results triggered discussion with national bodies such as the Association of Academic Coordinating Centers for Clinical Studies in Germany (KKS-network) or the Association of Medical Faculties/UMCs in Germany (MFT, Medizinischer Fakultätentag). At the same time, more attention was paid to the very low rate of summary results reporting in the EU Clinical Trials Register (EUCTR) across German (and many other European) UMCs for due trials [3]. Because these national discussions started in early 2019 first possible changes in the 5-years reporting rate can be assessed for trials completed in 2014 or later, which have not yet exceeded the 5-year period since completion. Changes in the 2-years reporting rate can only be assessed for trials completed in 2017 or later.

Our aim was to follow-up whether the 2- and 5-year reporting rates for German UMCs have increased over time and how UMCs differ in their contribution to timely and complete reporting. Here we report the results of a follow-up study for all trials from German UMCs that were registered on either ClinicalTrials.gov or DRKS and completed between 2014 and 2017.

## Methods

The study protocol was preregistered at https://osf.io/98j7u/.

This study applied the same tracking methods as in the former study. Broadly, we identified trials on ClinicalTrials.gov and DRKS using the following search criteria:

- Completion date between 2014-17 (per the registration on retrieval date)
- 35 German UMCs via their name in different spelling variants (ClinicalTrials.gov: UMC listed as responsible party, lead sponsor or with a principal investigator (PI) from UMC; DRKS: UMC listed as primary sponsor; Kiel and Lübeck share a single UMC since 2003; our current dataset represents this as a single UMC, whereas the former study included them as separate UMCs)
- Study type: interventional
- Study status: “Completed”, “Terminated”, “Suspended” or “Unknown” (or the equivalent DRKS categories). We include this additional criterion as anticipated completion dates can be entered in the registries, so non-completed trials can match the completion date criterion if the anticipated dates were not revised to reflect a status change.

We used the Aggregate Content of ClinicalTrials.gov (AACT) database [4] to retrieve a structured version of the ClinicalTrials.gov trial data, as well as additional metadata like phase, enrolment, and summary results deposition status and date. We manually searched and downloaded DRKS trials including all associated metadata from the registry website. Data was retrieved from both registries on June 3, 2020. The automatically detected UMC affiliations were then manually verified and studies that could not be clearly attributed to a German UMC were excluded from the sample.

For the verified German UMC trials, we performed a manual search for results publications (peer-reviewed journal article or dissertation) using the following search steps:

1. Identify the linked results publications on the ClinicalTrials.gov/DRKS website (both automatically and manually linked)
2. Search Google via trial ID (NCT ID or DRKS ID)
3. Search Google via other search parameters: official title, brief title, intervention name & PI

The publication search for a trial was stopped as soon as a peer-reviewed publication or dissertation matching the trial was found; Abstracts, posters and conference presentations were not counted as publications. The DOI/URL, publication date, and search step in which the publication was identified were recorded. If more than one results publication was identified in a search step, the earliest publication was recorded.

We assessed interrater reliability (IRR) across different search steps in a subset of 93 dual-searched trials, as prespecified in the study protocol, and predefined an agreement threshold of >85% (for details see methods supplement under https://osf.io/98j7u/). As the IRR for detection of results reporting fell below this threshold, an additional rater performed a second, independent manual search for all trials where no publication was found in the first round.

The retrieved metadata for the included trials from ClinicalTrials.gov and DRKS as well as the results from the manual publication search were combined with the results from the former study to form a single dataset. The dataset as well as a data dictionary is openly available from https://doi.org/10.5281/zenodo.5141343.

For the analysis of publication rates, we counted a trial as ‘timely’ published if either a publication or a summary result was published within two years after the completion date of the trial. A Kaplan-Meier curve visualizes the percentage of unpublished trials over time using the days from completion date to either publication or summary results reporting, using the R package KMsurv [5]. To account for the limited follow-up time of the publication search, the end date of the main publication search (September 1^st^, 2020) was used to calculate right censoring events in the estimation. For the calculation of UMC publication rates, a trial was counted for a UMC if it was mentioned as responsible party, lead sponsor or if it had a study PI from the UMC. UMCs only listed as study sites were excluded. If multiple UMCs fulfilled those criteria, the trial was counted for each UMC. We used Fisher’s exact test to compare the timely publication rates between this and the former study, and calculated an odds ratio as well as the 95% confidence intervals for each UMC. A positive odds ratio corresponds to a higher rate of timely publications in the second, more recent study.

We additionally assessed whether summary results reporting for the trials from the previous study period (2009-2013) has increased. For this, we retrieved the summary result status for all trials belonging to the earlier study cohort that were registered on ClinicalTrials.gov using the AACT version used in this study.

To identify variables that might have an influence on the timely publication of results, an exploratory logistic regression was used. To predict the timely publication status of all trials, the following explanatory variables were added to the model in a stepwise manner until no further model improvement could be observed: intervention type, mono/-multicentric, industry sponsor, completion year, recruitment status, phase, enrolment, center size. For a more detailed description of the data analysis, see the methods supplement under https://osf.io/98j7u/.

In addition, we compared our study’s findings for summary results reporting rates for each UMC in ClinicalTrials.gov and DRKS with that in EUCTR. For this, the matching names of the UMCs were identified on the EU Trials Tracker website [6], and the number of due and reported trials were retrieved from the GitHub repository of the project (https://github.com/ebmdatalab/euctr-tracker-data) for all considered UMCs in April 2021.

## Results

We identified 1,658 clinical trials via ClinicalTrials.gov (n=1,227) and DRKS (n=431) that i) were counted as lead trial for at least one German UMC and ii) had their completion date between 2014 and 2017. Of these trials, 402 (24%) investigated drugs while 276 (17%) investigated devices; the remaining trials were assigned to other intervention types (549 trials, 33%) or mentioned no intervention type (431 trials, 26%). In total, these trials included 371,173 anticipated participants. Table 1 presents more detailed trial characteristics including type of intervention, start date, and trial phase.

**Table 1:** Demographic data for all investigated lead trials.

|  | Trials |  |
| --- | --- | --- |
| Total | 1658 | 100% |
| Type of intervention |  |  |
| Behavioural | 138 | 8% |
| Biological | 45 | 3% |
| Combination Product | 1 | 0% |
| Device | 276 | 17% |
| Diagnostic Test | 1 | 0% |
| Dietary supplement | 45 | 3% |
| Drug | 402 | 24% |
| Genetic | 2 | 0% |
| Other | 159 | 10% |
| Procedure | 134 | 8% |
| Radiation | 24 | 1% |
| Not given | 431 | 26% |
| Lead sponsor<sup>1</sup> |  |  |
| Industry | 224 | 14% |
| Academia | 1434 | 86% |
| Phase |  |  |
| I | 83 | 5% |
| I-II | 50 | 3% |
| II | 234 | 14% |
| II-III | 26 | 2% |
| III | 136 | 8% |
| IV | 112 | 7% |
| Not given | 1017 | 61% |
| Mono-/Multicentric |  |  |
| Multicentric | 565 | 34% |
| Monocentric | 1093 | 66% |
| Number of participants |  |  |
| 1-99 | 1120 | 68% |
| 100-500 | 435 | 26% |
| >500 | 100 | 6% |
| Not given | 3 | 0% |
| Time of registration |  |  |
| Prospective registration<sup>2</sup> | 900 | 54% |
| After trial start | 756 | 46% |
| After trial completion | 210 | 13% |
| After publication | 25 | 2% |
| Start date not given | 2 | 0% |
| Trial completion year |  |  |
| 2014 | 378 | 23% |
| 2015 | 441 | 27% |
| 2016 | 409 | 25% |
| 2017 | 430 | 26% |
| Trial status |  |  |
| Completed | 1285 | 78% |
| Terminated | 144 | 9% |
| Suspended | 4 | 0% |
| Unknown status | 225 | 14% |

<sup>1</sup>Trials were counted for both an industry and UMC lead sponsor if both are listed as responsible party, lead sponsor or study PI.
<sup>2</sup>A registration counted as prospective registration if the study was registered in the month of the study start or earlier. This definition is necessary as start dates are often only specified to the month.

Out of all 1,658 trials, n=900 (54%) were registered prospectively (if the study was registered in the month of the study start or earlier), and 756 (46%) were registered after the reported start month (either actual or planned), including 210 (13%) registered after the completion date (Table 1). 100 trials (6%) included more than 500 anticipated participants. A total of 1,285 trials (78%) were completed and 148 (9%) were either terminated early or suspended; for 225 trials (14%), the status was unknown.

### Overall results reporting

For the former study [1], we developed an interactive website (https://s-quest.bihealth.org/intovalue/) that allows users to select and combine the measurement variables to benchmark all 36 German UMCs. This website now includes data from both studies (2009-2013 and 2014-2017), and can display results across both periods as well as for each individual period. In the following sections, we report key findings for the latest period (2014-2017) and changes in comparison to the previous study period (2009-2013).

Of all assessed trials in the present study, 43% (n=709) published their results in a timely manner, that is within 24 months after completion date. The dissemination route for timely reporting was by either journal publications only (37%, n=620), or summary results only (2%, n=33), or via both journal publications and summary results (2%, n=34), or dissertations (1%, n=22). At the level of German UMCs, the publication rate varied from 14% to 71% (Table 2).

**Table 2:**
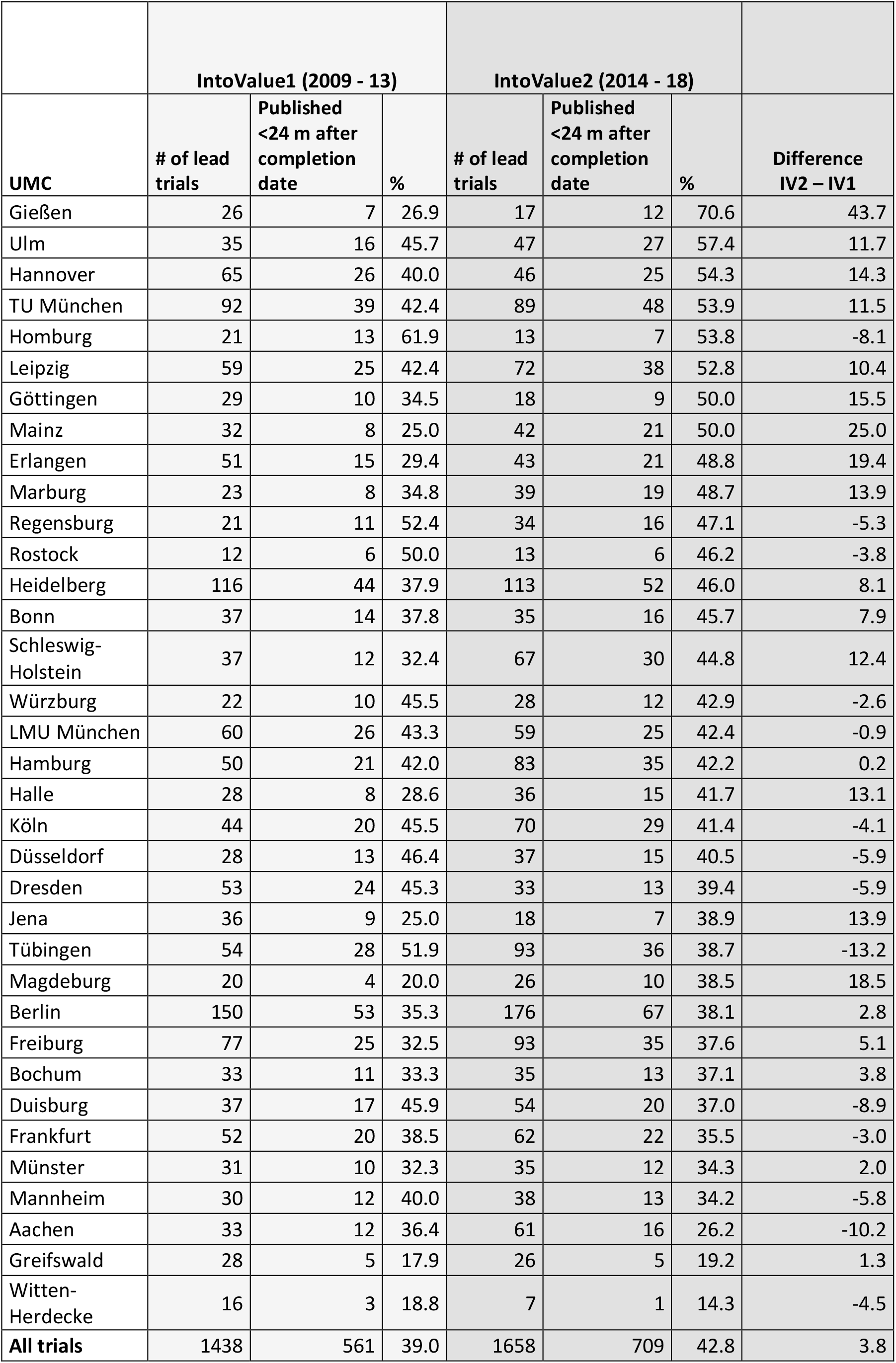
Timely publication rates at the level of individual German university medical centers (UMCs) and comparison to the timely publication rates of the former study.

|  | IntoValue1 (2009 - 13) |  |  | IntoValue2 (2014 - 18) |  |  |  |
| --- | --- | --- | --- | --- | --- | --- | --- |
| UMC | # of lead trials | Published <24 m after completion date | % | # of lead trials | Published <24 m after completion date | % | Difference IV2 – IV1 |
| Gießen | 26 | 7 | 26.9 | 17 | 12 | 70.6 | 43.7 |
| Ulm | 35 | 16 | 45.7 | 47 | 27 | 57.4 | 11.7 |
| Hannover | 65 | 26 | 40.0 | 46 | 25 | 54.3 | 14.3 |
| TU München | 92 | 39 | 42.4 | 89 | 48 | 53.9 | 11.5 |
| Homburg | 21 | 13 | 61.9 | 13 | 7 | 53.8 | -8.1 |
| Leipzig | 59 | 25 | 42.4 | 72 | 38 | 52.8 | 10.4 |
| Göttingen | 29 | 10 | 34.5 | 18 | 9 | 50.0 | 15.5 |
| Mainz | 32 | 8 | 25.0 | 42 | 21 | 50.0 | 25.0 |
| Erlangen | 51 | 15 | 29.4 | 43 | 21 | 48.8 | 19.4 |
| Marburg | 23 | 8 | 34.8 | 39 | 19 | 48.7 | 13.9 |
| Regensburg | 21 | 11 | 52.4 | 34 | 16 | 47.1 | -5.3 |
| Rostock | 12 | 6 | 50.0 | 13 | 6 | 46.2 | -3.8 |
| Heidelberg | 116 | 44 | 37.9 | 113 | 52 | 46.0 | 8.1 |
| Bonn | 37 | 14 | 37.8 | 35 | 16 | 45.7 | 7.9 |
| Schleswig-Holstein | 37 | 12 | 32.4 | 67 | 30 | 44.8 | 12.4 |
| Würzburg | 22 | 10 | 45.5 | 28 | 12 | 42.9 | -2.6 |
| LMU München | 60 | 26 | 43.3 | 59 | 25 | 42.4 | -0.9 |
| Hamburg | 50 | 21 | 42.0 | 83 | 35 | 42.2 | 0.2 |
| Halle | 28 | 8 | 28.6 | 36 | 15 | 41.7 | 13.1 |
| Köln | 44 | 20 | 45.5 | 70 | 29 | 41.4 | -4.1 |
| Düsseldorf | 28 | 13 | 46.4 | 37 | 15 | 40.5 | -5.9 |
| Dresden | 53 | 24 | 45.3 | 33 | 13 | 39.4 | -5.9 |
| Jena | 36 | 9 | 25.0 | 18 | 7 | 38.9 | 13.9 |
| Tübingen | 54 | 28 | 51.9 | 93 | 36 | 38.7 | -13.2 |
| Magdeburg | 20 | 4 | 20.0 | 26 | 10 | 38.5 | 18.5 |
| Berlin | 150 | 53 | 35.3 | 176 | 67 | 38.1 | 2.8 |
| Freiburg | 77 | 25 | 32.5 | 93 | 35 | 37.6 | 5.1 |
| Bochum | 33 | 11 | 33.3 | 35 | 13 | 37.1 | 3.8 |
| Duisburg | 37 | 17 | 45.9 | 54 | 20 | 37.0 | -8.9 |
| Frankfurt | 52 | 20 | 38.5 | 62 | 22 | 35.5 | -3.0 |
| Münster | 31 | 10 | 32.3 | 35 | 12 | 34.3 | 2.0 |
| Mannheim | 30 | 12 | 40.0 | 38 | 13 | 34.2 | -5.8 |
| Aachen | 33 | 12 | 36.4 | 61 | 16 | 26.2 | -10.2 |
| Greifswald | 28 | 5 | 17.9 | 26 | 5 | 19.2 | 1.3 |
| Witten-Herdecke | 16 | 3 | 18.8 | 7 | 1 | 14.3 | -4.5 |
| **All trials** | **1438** | **561** | **39.0** | **1658** | **709** | **42.8** | **3.8** |

In the former study, we found a steady improvement in timely publication from 35% for trials completed in 2009/2010, to 42% for trials completed in 2012/2013. This steady improvement did continue in the new sample but at a slower pace. Figure 1 presents the percentage of unpublished trials over time, stratified by completion year.

**Figure 1:**
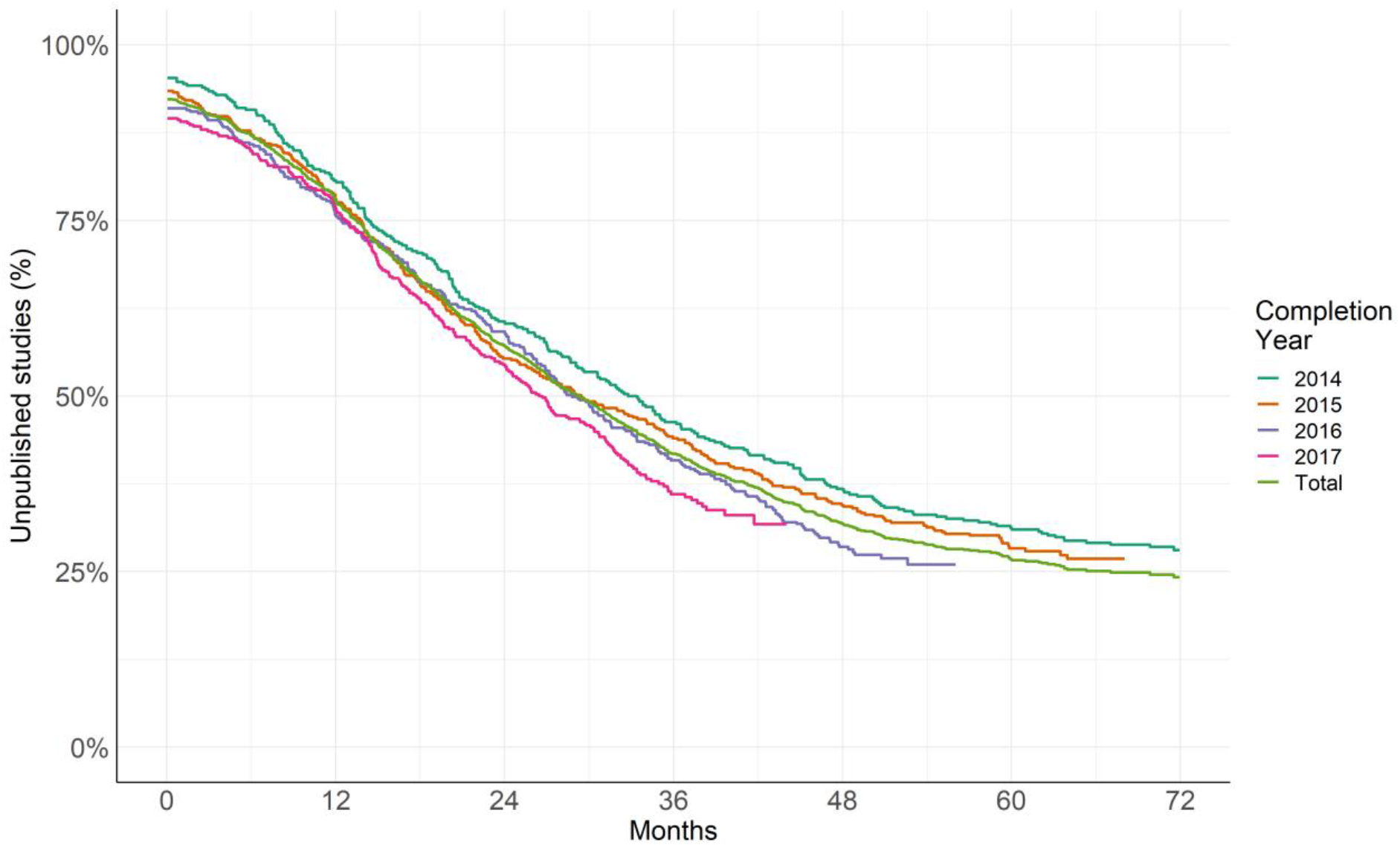
Kaplan-Meier curve showing the percentage of unpublished trials over time, stratified by completion year. A trial was counted as published if either a journal publication or summary result was found.

Table 2 compares the publication rates for the different UMCs of the former and the current study sample. The overall rate of trials published within 24 months is 3.8 percentage points higher than in the former study. The changes at the UMC-level varied from -13% to +44%. We found an improvement over time for the overall publication rates across all UMCs as well as for one individual UMC (Gießen); an odds ratio analysis comparing the publication rates for the current and the previous study showed that for those cases the confidence interval of the odds ratios did not include 1 (Figure 2).

**Figure 2:**
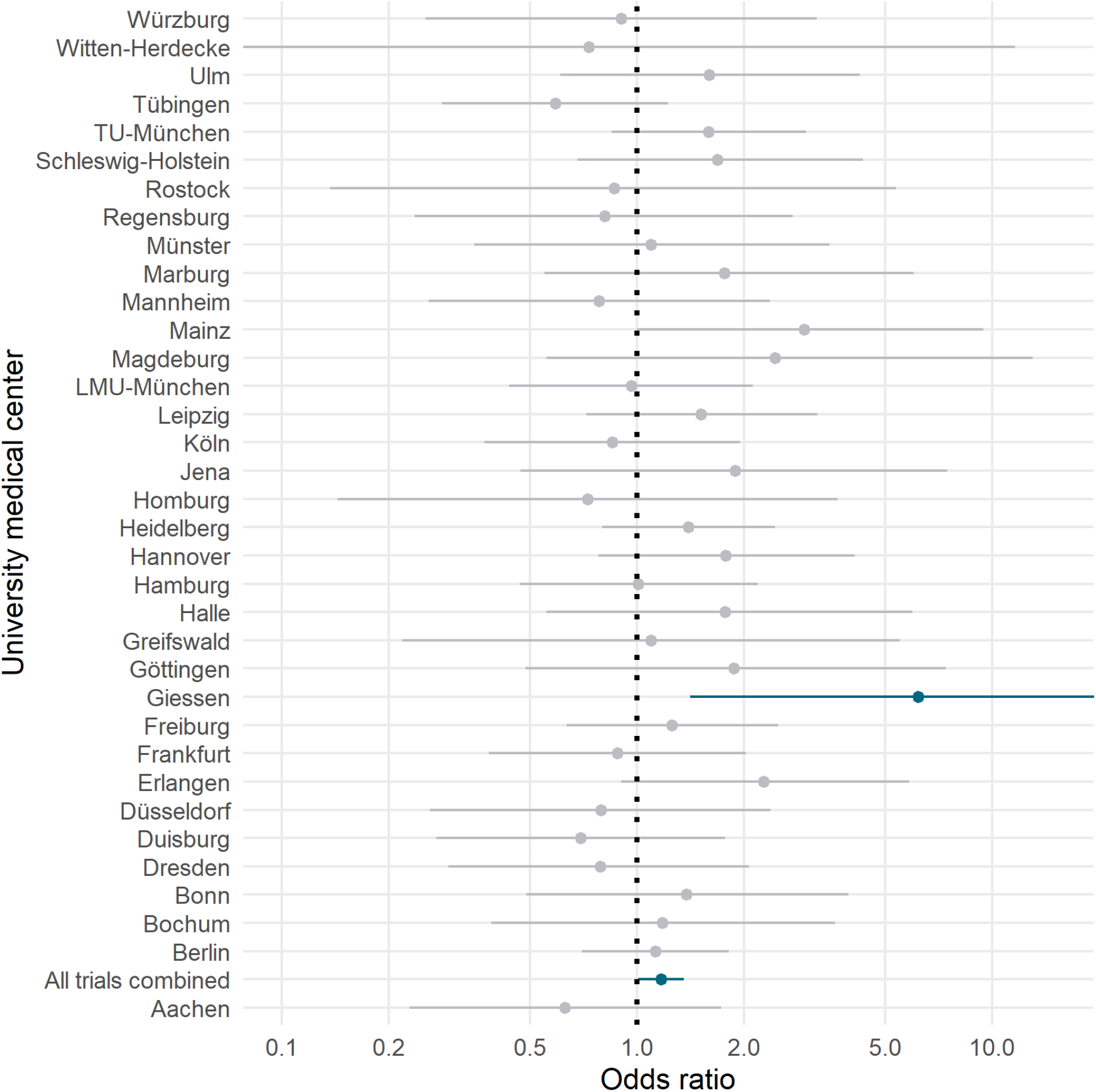
Odds ratios and 95% confidence intervals for the differences in timely publication rates (within 24 months) between the first and the second study. For each UMC, Fisher’s exact test was performed comparing the numbers of trials with and without timely publication for the two studies. Here, a positive odds ratio corresponds to a higher rate of timely publications in the second, more recent study.

Of the 1,658 trials, there was a subgroup of 635 trials where the observation period from completion date to our publication search was at least five years. For this subgroup, we found that 70% published their results within five years varying from 50% to 100% across UMCs. The proportion of trials published within five years was 67% in the sample of the first study (n=959).

Only 30 (2.4%) of the trials registered on ClinicalTrials.gov reported summary results within 12 months after completion, increasing to 62 (5.1%) of 1,227 trials within 24 months. This is similar to the 4.7% of trials reporting summary results after 24 months observed in the former study. Reanalysis of the trials from the earlier study using the newly downloaded AACT data showed that when any follow-up time was considered (cutoff date: June 3, 2020), summary results were found for 105 (8.4%) of all 1,243 lead trials, a minor increase from the 91 (7.3%) trials with summary results detected in the earlier study.

It is important to note that German drug trials are required by law to report summary results in the EUCTR within 12 months after completion. In April 2021, summary results reporting for due trials from German UMCs in EUCTR was 39% in average according to the EU TrialsTracker [6] with a variation of 0-100% across all German UMCs (see supplemental Table S1).

Altogether, 21,294 participants were planned to be included in the 188 trials that did not publish their results within five years after completion. Extrapolated to the full sample of lead trials (21,294 × 1658 ÷ 635 ÷ 4), an average of 13,900 planned participants per year were enrolled in trials from German UMCs that did not disseminate their results within five years after completion.

### Subgroup analyses

To identify subgroups with substantially different publication rates, we performed an additional exploratory logistic regression analysis (see supplementary file under https://osf.io/98j7u/ for more details). ‘Study status’ (completed: 47% timely reported trials after 24 months, terminated/suspended/unknown: 27%) and ‘Mono-/Multicentric’ (multicentric: 47%, monocentric: 40%) had the strongest associations with publication rates. However, these associations were weaker compared with the former study and thus were not sufficient to describe which studies were reported in time with great confidence.

## Discussion

This study found that of all 1,658 clinical trials led by one of 35 German UMCs registered on ClinicalTrials.gov or the German registry DRKS and completed between 2014 and 2017, only 43% disseminated results within 2 years after completion. Even five years after completion, 30% still did not disseminate any results via journal publications or via summary results in the two registries. Both, 2-year and 5-year publication rates show only slight improvements (4 and 3 percentage points higher, respectively) compared to our previous study on results reporting of clinical trials from German UMCs completed between 2009 and 2013 [1].

UMCs (as sponsors) and clinical trialists (as PIs) should launch a concerted effort to rectify past ethics violations by retrospectively making public the results of all their unreported trials. In addition, UMCs should put systems into place that ensure that all future trial results are made public on trial registries within 12 months of trial completion, as per WHO best practices. Results dissemination should be understood as a non-negotiable issue in the same way we understand ethics approval or informed consent. The risks and burdens for trial participants and the invested public resources (financial and personnel) are ethically justified only against the prospect of a community-wide knowledge gain. If results are not disseminated, this knowledge gain is not reached and the trial becomes unethical as no social value [7,8] is created to counterbalance the individual risks of the study participants.

There are two accepted measures for timeliness of results reporting: The joint statement of the World Health Organization (WHO) recommends reporting of summary results, which report the key results in tabular or summarized form, within 12 months, and as journal publication within 24 months [9]. Beside this WHO recommendation, European law since 2014 requires reporting of summary results for all drug trials in the EUCTR within 12 months after study completion [10]. German and other European UMCs improved their summary results reporting for drug trials in the EUCTR over the past two years, likely pushed by academic initiatives such as the EU TrialsTracker [6], NGOs such as Cochrane, Transparency International, TranspariMED [11,12], and media coverage [13].

Outside the world of regulated drug trials (representing 24% of all 1,658 clinical trials), the primary route for results dissemination remains publication in peer-reviewed journals. In line with WHO recommendations [9], UMCs and funders should require PIs to employ summary results as an effective and efficient way for timely results dissemination in all areas of clinical research. Furthermore, if results publications are delayed because of time-consuming peer-review processes, UMCs and funders could emphasize the publication of preprints as another way to increase the timeliness of results dissemination. The steep increase in preprints on COVID-19 research demonstrates the acceptance of preprints as a measure to disseminate research [14]. As many COVID-19-related trials were started in 2020 and as the pandemic has potentially limited the attention to non-COVID trials [15], a negative effect of the pandemic on trial reporting cannot be excluded.

There are several limitations to our approach. Despite an extensive search protocol, the manual searches might have missed results publications. However, results publications not found by such an extensive search protocol are likely inadequately linked to registered trials. Additionally, we relied on registry entries, for example regarding expected completion dates or cross-registration of clinical trials, which might be incomplete and outdated in several cases.

The current study involved major personnel resources to manually follow-up hundreds of clinical trials (approx. 24 person-days assuming 5 min. per trial and including the dual-searched trials). Only the summary results status of the trials was retrieved via automated procedures. The automated tracking of results reporting would be more efficient if authors link the available results publications at the registry website and if the publications include the respective trial IDs in meta-data, abstract, and full-text, as endorsed by CONSORT [16] and the WHO [9]. We are conducting a follow-up study on this trial sample to investigate linkage between registrations and publications.

Our study protocol and a detailed methods supplement (https://osf.io/98j7u/) allows the replication of our study also for other countries. A first replication based on the study protocol for our former study is already available for Poland [17].

The dataset and the accompanying website (https://s-quest.bihealth.org/intovalue/) allow UMCs to identify individual studies with overdue reporting, and we hope it will support UMCs to retrospectively make public missing trial results. We plan to re-assess UMCs’ progress in a future study.

## Data Availability

The dataset underlying the results is available from https://doi.org/10.5281/zenodo.5141343 under a CC BY 4.0 license. All analysis scripts used for this study are available from https://github.com/quest-bih/IntoValue2 under a MIT license.

https://github.com/quest-bih/IntoValue2

https://osf.io/98j7u/

## Acknowledgements

We would like to thank Ulf Tölch for providing fruitful ideas regarding the visualization of differences in publication rates over time.

## Contributorship statement

DS, NR and SW designed the study. NR, SW, TB, MH, HK, and EN performed the search. NR and MS performed the statistical analysis. All authors were involved in writing and editing the manuscript.

## Data Availability

The dataset underlying the results is available from https://doi.org/10.5281/zenodo.5141343 under a CC BY 4.0 license.

## Code availability

All analysis scripts used for this study are available from https://github.com/quest-bih/IntoValue2 under a MIT license.

