## Supplemental Table S1 for "Results dissemination from completed clinical trials conducted at German university medical centers remains delayed and incomplete. The 2014-2017 cohort"

Supplemental Table 1: Summary results reporting for due trials from German UMCs in EUCTR according to the EU Trials Tracker. Data was retrieved in April 2021. For the UMCs Greifswald and Witten/Herdecke no due trials were reported. For Mannheim, no matching entry was found.

| **Cities** | **EU Trials Tracker name** | **Percentage reported** | **Due trials** | **Reported trials** |
|---|---|---|---|---|
| Aachen | RWTH Aachen University | 13.3 | 15 | 2 |
| Berlin | Charité-Universitätsmedizin Berlin | 58.4 | 77 | 45 |
| Bochum | Ruhr University Bochum | 0 | 4 | 0 |
| Bonn | University of Bonn | 0 | 14 | 0 |
| Dresden | Dresden University of Technology | 76.2 | 21 | 16 |
| Duisburg | University Duisburg-Essen | 7.7 | 13 | 1 |
| Düsseldorf | Heinrich Heine University Düsseldorf | 80 | 5 | 4 |
| Erlangen | University Erlangen-Nuremberg | 58.3 | 24 | 14 |
| Frankfurt | Goethe University | 9.1 | 11 | 1 |
| Freiburg | University of Freiburg | 75.0 | 20 | 15 |
| Gießen | University Hospital Giessen and Marburg | 0 | 2 | 0 |
| Göttingen | University of Göttingen | 33.3 | 15 | 5 |
| Greifswald | Medical University Greifswald | - | 0 | 0 |
| Halle | Martin Luther University Halle-Wittenberg | 8.3 | 12 | 1 |
| Hamburg | University of Hamburg | 47.4 | 19 | 9 |
| Hannover | Hannover Medical School | 20.7 | 29 | 6 |
| Heidelberg | Heidelberg University Hospital | 65.5 | 29 | 19 |
| Homburg | Saarland University | 0 | 2 | 0 |
| Jena | Friedrich Schiller University Jena | 50.0 | 8 | 4 |
| Köln | University of Cologne | 77.8 | 36 | 28 |
| Leipzig | Leipzig University | 92.0 | 25 | 23 |
| LMU München | University of Munich (Ludwig-Maximilians) | 13.5 | 37 | 5 |
| Magdeburg | Otto von Guericke University Magdeburg | 100 | 9 | 9 |
| Mainz | Johannes Gutenberg University of Mainz | 44.8 | 29 | 13 |
| Mannheim | - | - | - | - |
| Marburg | Philipps-University Marburg | 57.1 | 7 | 4 |
| Münster | University of Münster | 77.3 | 22 | 17 |
| Regensburg | University of Regensburg | 22.2 | 9 | 2 |
| Rostock | Universität Rostock | 0 | 1 | 0 |
| Schleswig-Holstein | Schleswig-Holstein University Hospital | 0 | 9 | 0 |
| TU München | Technical University of Munich | 90.3 | 31 | 28 |
| Tübingen | University Hospital Tübingen | 17.6 | 17 | 3 |
| Ulm | University of Ulm | 53.3 | 15 | 8 |
| Witten-Herdecke | University of Witten/Herdecke | - | 0 | 0 |
| Würzburg | Julius Maximilian University of Würzburg | 12.5 | 8 | 1 |
